# A randomized clinical trial to stimulate the cholinergic anti-inflammatory pathway in patients with moderate COVID-19-pneumonia using a slow-paced breathing technique

**DOI:** 10.1101/2021.12.03.21266946

**Authors:** Elisabeth Maria Balint, Beate Grüner, Sophia Haase, Mandakini Kaw-Geppert, Julian F. Thayer, Harald Gündel, Marc N. Jarczok

## Abstract

**Importance:** Vagus nerve stimulation via slow-paced breathing could serve as an adjuvant therapeutic approach to reduce excessive inflammation in coronavirus disease 2019 (COVID-19) pneumonia.

**Objective:** Does a slow-paced breathing technique increasing vagal activity reduce Interleukin-6 (IL-6) in patients hospitalized with moderate COVID-19 pneumonia compared to standard care?

**Design:** Single-center randomized controlled clinical trial with enrolment from February 23^rd^ 2021 through June 17^th^ 2021 and follow-up until July 22^nd^ 2021.

**Setting:** Ward for infectious diseases and temporary COVID-19 ward, Ulm University Hospital, Germany

**Participants:** Consecutive sample of patients hospitalized with confirmed severe acute respiratory syndrome coronavirus 2 (SARS-CoV-2) infection and moderate COVID-19 pneumonia (primary diagnosis). Of 131 patients screened, 48 patients were randomized and 46 patients analyzed (N=23 per group).

**Interventions:** Slow-paced 20-minute breathing exercise three times a day with six breaths per minute (inhalation-to-exhalation ratio 4:6).

**Main outcomes and measures:** Differences between intervention and control group in IL-6 calculated using multilevel mixed-effect linear regression models with random slope including the covariates relevant comorbidities, COVID-19 medication, and age.

**Results:** Mean age 57 years±13 years, N= 28 (60%) male, N=30 (65%) with relevant comorbidities.

The model including group by time interaction revealed a significantly lower trajectory of IL-6 in the intervention group compared to the control group (effect size Cohens f^2^=0.11, LR-test p=.040) in the intention-to-treat sample, confirmed by treatment-per-protocol analysis (f^2^=0.15, LR-test p=.022). Exploratory analysis using the median split of practice time to predict IL-6 of the next morning indicated a dose-response relationship with beneficial effects of practice time above 45 minutes a day.

Three patients in each group were admitted to ICU, one died. Oxygen saturation increased during slow-paced breathing (from 95.1%±2.1% to 95.4%±1.6%, p=0.006).

**Conclusion and relevance:** Patients practicing slow-paced breathing had significantly lower IL-6 values than controls with a small to medium effect size and without relevant side effects. Further trials should evaluate clinical outcomes as well as an earlier start of the intervention, i.e., at symptom onset. This would offer an access to a therapy option not only for high-income, but also for low- and middle-income countries.

**Trial registration:** German register of clinical trials (ID: DRKS00023971) https://www.drks.de, Universal Trial Number (UTN) U1111-1263-8658;

## Introduction

The pandemic of severe acute respiratory syndrome coronarvirus 2 (SARS CoV2) still occupies politics and health care. Though immunization is available now and knowledge about therapy options has widely progressed, there is still an urgent need especially for cheap, broadly accessible interventions that could be spread globally.

A characteristic problem occuring in coronavirus disease 2019 (COVID-19) are excessive elevations of pro-inflammatory cytokines such as interleukin-6 (IL-6) and C-reactive protein (CRP) which are associated with worse clinical courses (1,2). Several trials have tested anti-inflammatory agents, including dexamethasone or IL-6 antagonists with beneficial outcomes (3–5). Beside drugs, there exists a clinically relevant, non-pharmaceutical way to reduce inflammation through vagus nerve stimulation (VNS). The latter is involved in the regulation of the immune response via the cholinergic anti-inflammatory reflex (6). Specifically, the efferent vagally mediated reflex arc regulates inflammation and the release of pro-inflammatory cytokines such as IL-6 from acetylcholine-synthesizing T-cells (7). Accordingly, plasma levels of pro-inflammatory cytokines increase with cervical or subdiaphragmatic vagotomy, whereas electrical VNS or acetylcholine administration decrease IL-6 cytokine levels in human and mouse models (7). Measures of vagal activity and inflammatory parameters such as IL-6, CRP, and tumor necrosis factor α (TNF-α) are strongly correlated (8). In the specific case of infection with SARS-CoV-2, as the cholinergic anti-inflammatory mechanism controls NF-κB action through Acetylcholine coupled to the α7n-Acetylcholin-receptor (9), insufficient vagal activity appears to be the cause of both unhindered viral replication and uncontrolled cytokine release along the virus-driven NF-κB pathway (10). Therefore, increasing vagal activity seems to be a promising therapeutic approach.

The vagus nerve can be stimulated with electric devices (11,12). The authors of a randomized controlled study proved feasibility for auricular electrical VNS in 31 patients hospitalized due to COVID-19 and did not report side effects, but were not able to show clinical improvement (13). Two case reports describe decreasing IL-6 and CRP after onset of electrical VNS in four COVID-19 patients suffering from moderate or severe COVID-19-pneumonia (14,15). However, electrical VNS has the limitation that no device has yet been certified for this anti-inflammatory use, this intervention must be performed by medical professionals, and is therefore limited with respect to personal and financial resources. To our knowledge, no study investigated the effect of non-electrical VNS in patients with COVID-19. A simple way to increase the activity of the vagus nerve is a specific breathing technique with reduced frequency and a prolonged exhalation phase (slow-paced breathing) (16,17). Though some studies show effects of slow-paced breathing on IL-6, CRP or TNF-α after several days or weeks in patients with hypertension (18) and irritable bowel syndrome (19), the effect of slow-paced breathing in infectious diseases was not yet investigated.

Therefore, we performed a randomized controlled trial in patients with moderate COVID-19 pneumonia to investigate the hypothesis whether a breathing technique that increases vagal activity reduces inflammatory levels (primary outcome: IL-6; secondary outcome: CRP, leukocytes) in patients with COVID -19 pneumonia compared to a control group of patients with treatment as usual.

## Materials and methods

The present study design is a prospective, two-arm, open-label, single-center randomized controlled trial. The protocol was approved by the Institutional Review Board of Ulm University (No. 3/21, 01/02/2021) and registered prior to screening start at the German Clinical Trials Register (ID: DRKS00023971), Universal Trial Number (UTN) U1111-1263-8658. The study was conducted in compliance with the Declaration of Helsinki, the Guideline for Good Clinical Practice, and local regulatory requirements. All patients provided written informed consent prior to inclusion.

### Patients

Patients enrolled were hospitalized with SARS-CoV-2-infection confirmed by positive polymerase chain reaction assay and the primary diagnosis of a moderate degree of COVID-19 according to WHO definition. The following exclusion criteria were applied: 1) Severe COVID-19 pneumonia with fever and bilateral lung infiltrates and respiratory rate>30/min or SpO2<90% on room air (adapted from WHO), or 2) condition after surgery/trauma/acute event (stroke, myocardial infarction, acute COVID-independent infection) in the last four weeks, i.e., other primary diagnoses than COVID-19, or 3) current pregnancy, or 4) patients with pre-existing pulmonary disease who were on oxygen prior to infection (e.g., due to pulmonary fibrosis, COPD), or 5) limited ability to give consent (e.g., due to dementia), or 6) limited ability to perform breathing maneuvers independently (e.g., high frailty), or 7) limited ability to provide self-care (German care level two or three), or 8) insufficient language skills, or 9) seizures in the medical history.

### Study design

Screening was performed starting from 23/02/2021 until 17/06/2021 on either the ward for infectious diseases or the temporary ward for patients suffering from COVID-19 at the Clinic for Internal Medicine III (Infectious Diseases, hematology, oncology) of the University Medical Center Ulm (Germany). Consenting patients were randomized in a 1:1 ratio to receive the breathing intervention (intervention group, IG) or standard care (control group, CG). The randomization list was created prior to screening start by MNJ with blocks of 20, 16, 14, and 16 numbers to account for the adaptive design (see below Statistical methods section) using the software STATA (20) and was not accessible to recruiting study personnel (EMB, SH, MKG).

Patients randomized to intervention group were asked to perform the 20-minute breathing exercise three times a day with six breaths per minute and an inhalation to exhalation ratio of 4:6 seconds (see Supplement for the instruction). To support the correct technique, the free application BreathBall (https://breathball.com/de/home-de/) was shown on a smartphone. The application facilitates paced breathing by displaying a decreasing (exhale) and increasing (inhale) ball, combined with sound if preferred. The study personnel monitored the first exercise to assist and register side effects. If the patients were unable to follow the breathing scheme, it was adapted slightly up to a breathing frequency of 10 per minute. Further exercises were to be done independently and the time spent in the exercise was self-recorded. The control group received standard care. Both groups were visited every second day by the study personnel to assess symptoms, check for side effects, record the practice times and to collect oxygen saturation, oxygen flow and breathing frequency. Blood samples were taken in routine at approximately 8am in the morning and analyzed by the clinical chemistry within four hours by accredited procedures to assess IL-6, CRP, and leucocytes and tumor-necrosis-factor alpha (TNF-α).

Four weeks after discharge, patients were contacted via telephone to assess symptoms and adverse events (follow-up). Follow-up was completed on July 22^nd^, 2021.

### Measures

Only one blood sample measurement was included per day. If several blood samples were available, we included the measurement closest to 8am. Values below the detection limit (IL6<1.5 pg/ml, CRP < 0.6 mg/l, TNF-α < 8.1 pg/ml) were set to the value of the detection limit.

Breathing protocol adherence was defined as follows: if the breathing intervention was performed at least once autonomously and if the percentage of minutes in paced breathing were at least 50% of the required minutes, i.e., min. 30 minutes per day averaged over the whole stay. Treatment Per Protocol Sample (TPP) included only those patients meeting the adherence criteria.

Adverse events were defined as transfer to ICU or death.

Screening for depressive and anxious symptoms was performed using the PHQ-4 (positive screening if sum score ≥ 3) (21).

### Statistical analysis

The Institutional Review Board requested an adaptive design (power of 80% and an alpha of .05). After N=30, N=46 and N=60 of patients, the effect size can be calculated. The intervention can be stopped at N=46 if an effect size of greater than f=0.16 exists. After N=46, data were reviewed resulting in a significant effect size of f=.11 (ITT) and f=.14 (TPP). Considering the given seasonal circumstances (infection rate lowering, no further patient admissions) the study was discontinued mid of June 2021 to avoid delay (new patient admissions expected only five months later) and to limit the sample to one wave.

### Statistics

For comparisons between the intervention and control group, chi square tests and the Mann-Whitney U test were used if appropriate. For repeated measurements (oxygen saturation, oxygen flow, breathing frequency), multilevel-mixed effect linear regression models were calculated.

Due to a skewed distribution, IL-6, TNF-α, leucocytes, and CRP were natural log-transformed prior to parametric statistical testing to better approximate Gaussian distribution. The level of significance was set a priori to p<.05 (two-sided). Data management and analysis were performed using STATA 15.1 SE (STATA Corp., College Station, Texas, USA).

#### Trial outcome analysis method

Per outcome, four multilevel mixed-effect linear regression models were calculated and compared, as recommended by a recently published best practice guidance for linear mixed-effects models. (22) The covariance was set to unstructured. The first model included random effect only (on the individual level), the second additionally included the main fixed effects for group (IG vs. CG) and time (days since study inclusion). Since clinically meaningful differences existed between the study groups (see Table 1) additional covariates were also included. These were: relevant comorbidities (no vs. yes), COVID-19 pneumonia medication (count), and age (years). The third model additionally included the variable time to the random effect equation. The fourth model additionally included the interaction between group and time in the fixed effect part. The model fit was compared between these four models and parsimonious model improvement was tested using likelihood ratio tests (see Table 2). Additionally, information criteria (Akaike IC and Bayesian IC) were assessed.

**Table 1:**
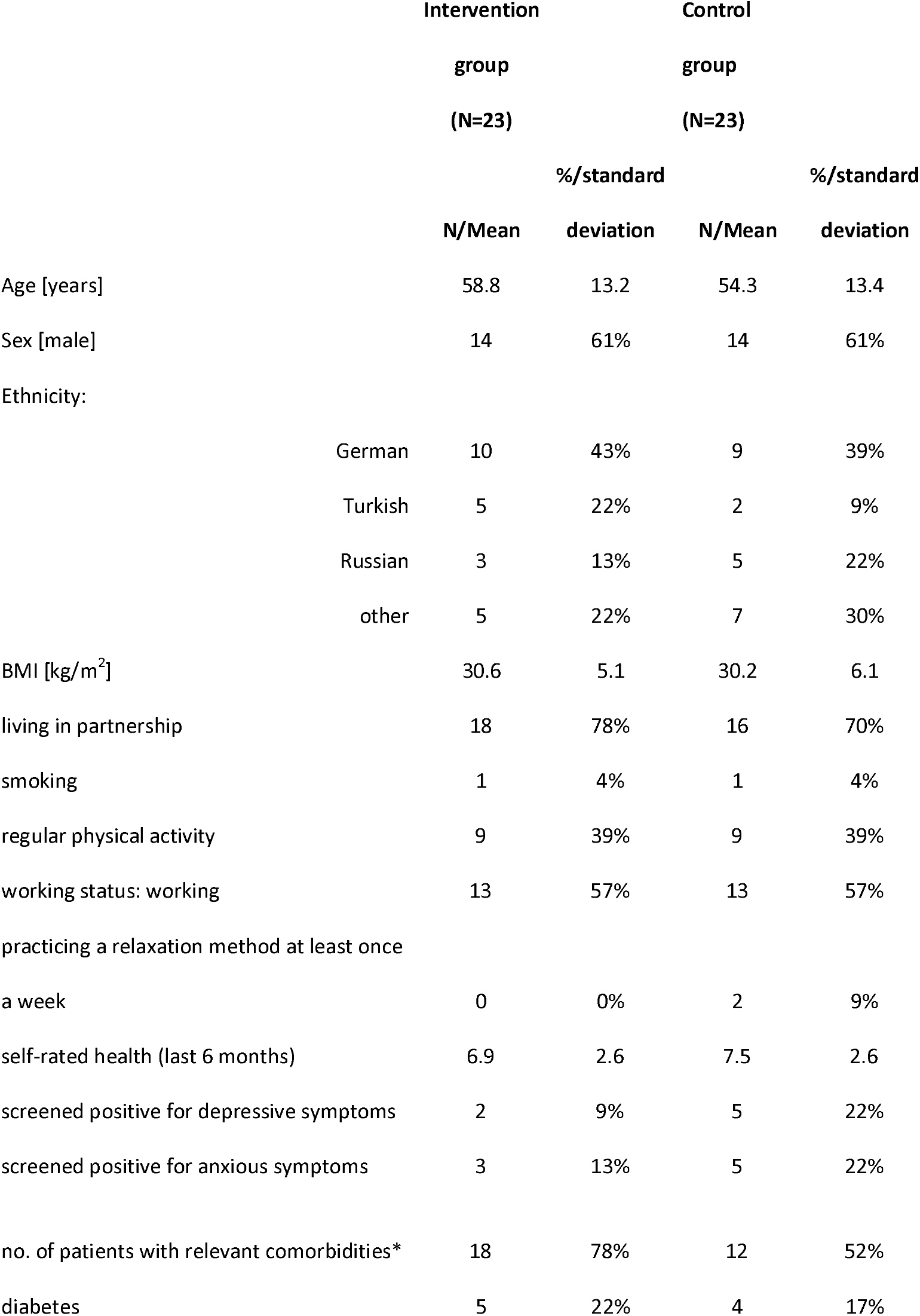
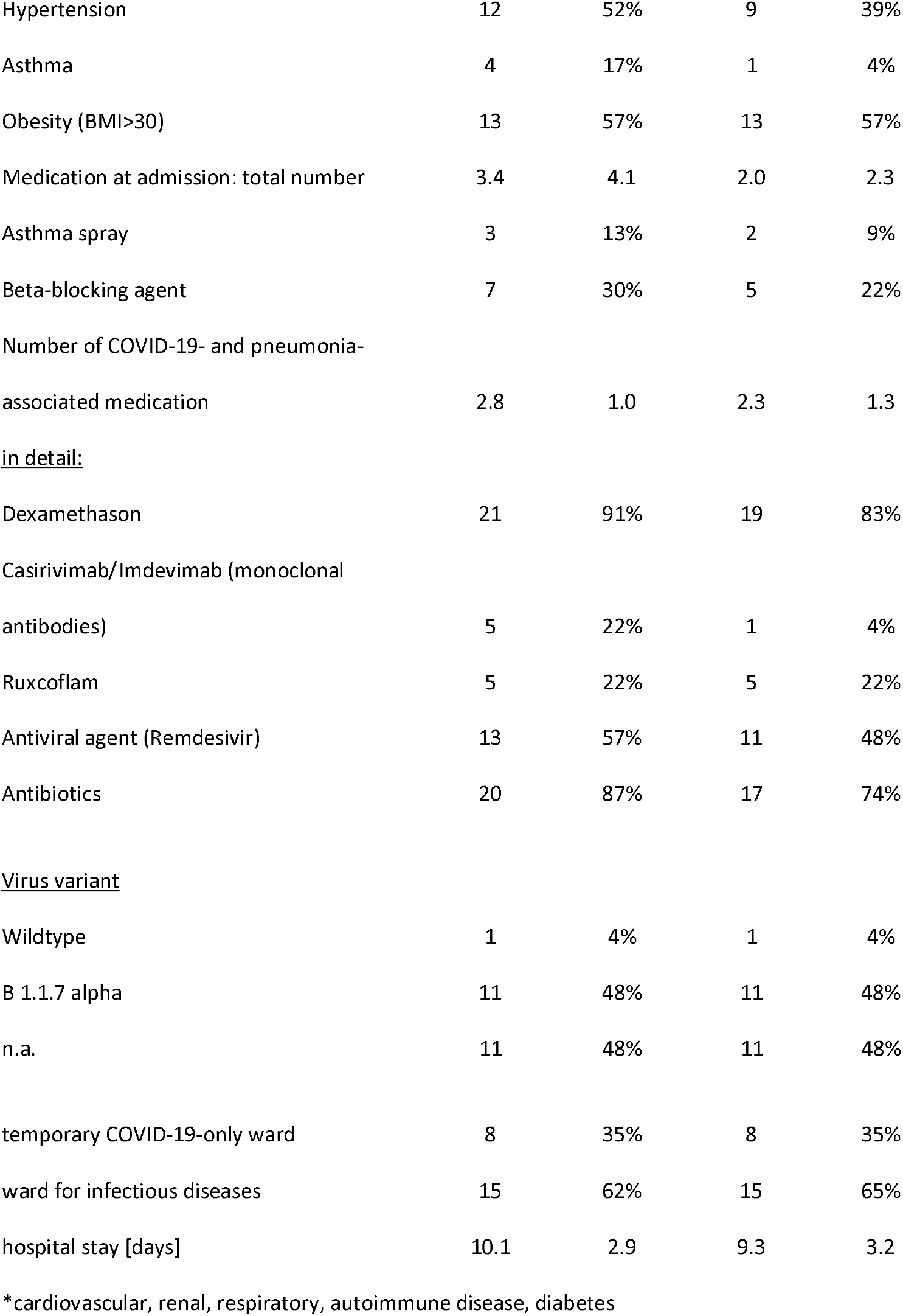

**Table 2:** Model comparison of ITT analysis by outcome (N=46). **BOLD** lines indicate favored model

| DV | Obs | Model Number | Model specification | Fixed Effects added | Random effects |  | Model fit |  |  | LRT Test against nested |  |  |  |
| --- | --- | --- | --- | --- | --- | --- | --- | --- | --- | --- | --- | --- | --- |
|  |  |  |  |  | Subjects (ID) | Item (Day) | AIC | BIC | LL | df | df | X2 | Prob > chi2 |
| IL-6<br>(ln[pg/ml]) | 208 (min. 2, avg 4.5, max 10) | 1 | RE only | - | intercepts | - | 684.1391 | 694.1518 | -339.06957 | 3 | - | - | - |
|  |  | 2 | M1 + FE main effects | Group + Day | intercepts | - | 682.5409 | 749.2917 | -321.2705 | 20 | 17 | 35.60 | 0.0052 |
|  |  | 3 | M2 + RE | - | intercepts | intercepts | 674.3049 | 747.7307 | -315.1524 | 22 | 2 | 12.24 | 0.0022 |
|  |  | **4** | **M3 + Interaction** | **Group X Time** | **intercepts** | **intercepts** | **677.1227** | **793.9366** | **-303.5614** | **35** | **13** | **23.18** | **0.0395** |
| Leucocytes<br>(ln[giga/l]) | 214 (min. 2, avg 4.7, max 10) | 1 | RE only | - | intercepts | - | 195.8852 | 205.9832 | -94.94262 | 3 | - | - | - |
|  |  | 2 | M1 + FE main effects | Group + Day | intercepts | - | 127.7382 | 195.0577 | -43.86911 | 20 | 17 | 102.15 | <0.0001 |
|  |  | 3 | **M2 + RE** | - | **intercepts** | **intercepts** | **111.1439** | **185.1954** | **-33.57196** | **22** | **2** | **20.59** | **<0.0001** |
|  |  | 4 | M3 + Interaction | Group X Time | intercepts | intercepts | 124.6047 | 239.0478 | -28.30233 | 34 | 12 | 10.54 | 0.5688 |
| CRP<br>(ln[mg/l]) | 222 (min. 2, avg 4.8, max 10) | 1 | RE only | - | intercepts | - | 718.8937 | 729.1018 | -356.4469 | 3 | - | - | - |
|  |  | 2 | M1 + FE main effects | Group + Day | intercepts | - | 589.4273 | 657.4808 | -274.7136 | 20 | 17 | 163.47 | <0.0001 |
|  |  | 3 | **M2 + RE** | - | **intercepts** | **intercepts** | **504.5118** | **579.3707** | **-230.2559** | **22** | **2** | **88.92** | **<0.0001** |
|  |  | 4 | M3 + Interaction | Group X Time | intercepts | intercepts | 516.4878 | 635.5815 | -223.2439 | 35 | 13 | 14.02 | 0.3722 |
| TNF-α<br>(ln[pg/ml]) | 151 (min. 1, avg 3.4, max 8) | 1 | RE only | - | intercepts | - | 189.4543 | 198.5061 | -91.72714 | 3 | - | - | - |
|  |  | 2 | M1 + FE main effects | Group + Day | intercepts | - | 182.9308 | 243.2764 | -71.46538 | 20 | 17 | 40.52 | 0.0011 |
|  |  | 3 | **M2 + RE** | - | **intercepts** | **intercepts** | **170.7575** | **237.1376** | **-63.37873** | **22** | **2** | **16.17** | **0.0003** |
|  |  | 4 | M3 + Interaction | Group X Time | intercepts | intercepts | 175.1232 | 274.6934 | -54.5616 | 33 | 11 | 17.63 | 0.0905 |

These models were calculated for each outcome and each analysis sample (ITT and TPP). The analyses were restricted to a maximum of thirteen days after study inclusion, because afterwards no observations were available in the IG. This led to a deletion of three observations from three patients of the CG.

### Post hoc analyses

Potential dose-response effects from categorized breathing minutes on daily IL-6 values were analyzed in all patients from the intervention group. Daily total breathing minutes were dichotomized at median value (45 minutes). Categorized minutes of breathing were related to blood samples from the following morning to retain temporal relationship. Two multilevel mixed-effect linear regression models were calculated for the primary outcome IL-6. The first model included the categorized practice time in the fixed effect part. The random effect part included the individual slopes as well as the binary practice time as cross-level interaction. Findings for analyses of end points other than the primary end point should be interpreted as exploratory due to the potential for type I error using multiple comparisons.

## Results

### Study population

Of 131 patients screened, 81 met exclusion criteria (see Figure 1). Main exclusion reasons were invasive procedures, trauma or acute myocardial infarction/stroke during the last four weeks (N=35, 43%), followed by high frailty and dementia (N=23, 28%) and insufficient knowledge of German language (N=16, 20%). Out of the remaining 50 patients, two patients (4%) were not willing to participate. A total of 48 patients (37% out of 131 patients screened) were randomized. Monitoring during the study revealed an exclusion criterion in two patients (severe COVID-19 pneumonia before study entry). Therefore, 46 patients (N=23 patients per group) were available for intention to treat (ITT) analysis. Seven patients in the intervention group practiced less than 50%. Two stopped because they had difficulties with the implementation of the breathing exercise in terms of concentration and technique. Two were transferred to ICU within two days of study entry due to deterioration of COVID-19 pneumonia. Three practiced continuously, but with shorter duration or less frequently, in total less than 50% of the required time. Thus, 16 patients of the IG and 23 of the CG entered the treatment per protocol (TPP) analysis.

**Figure 1:**
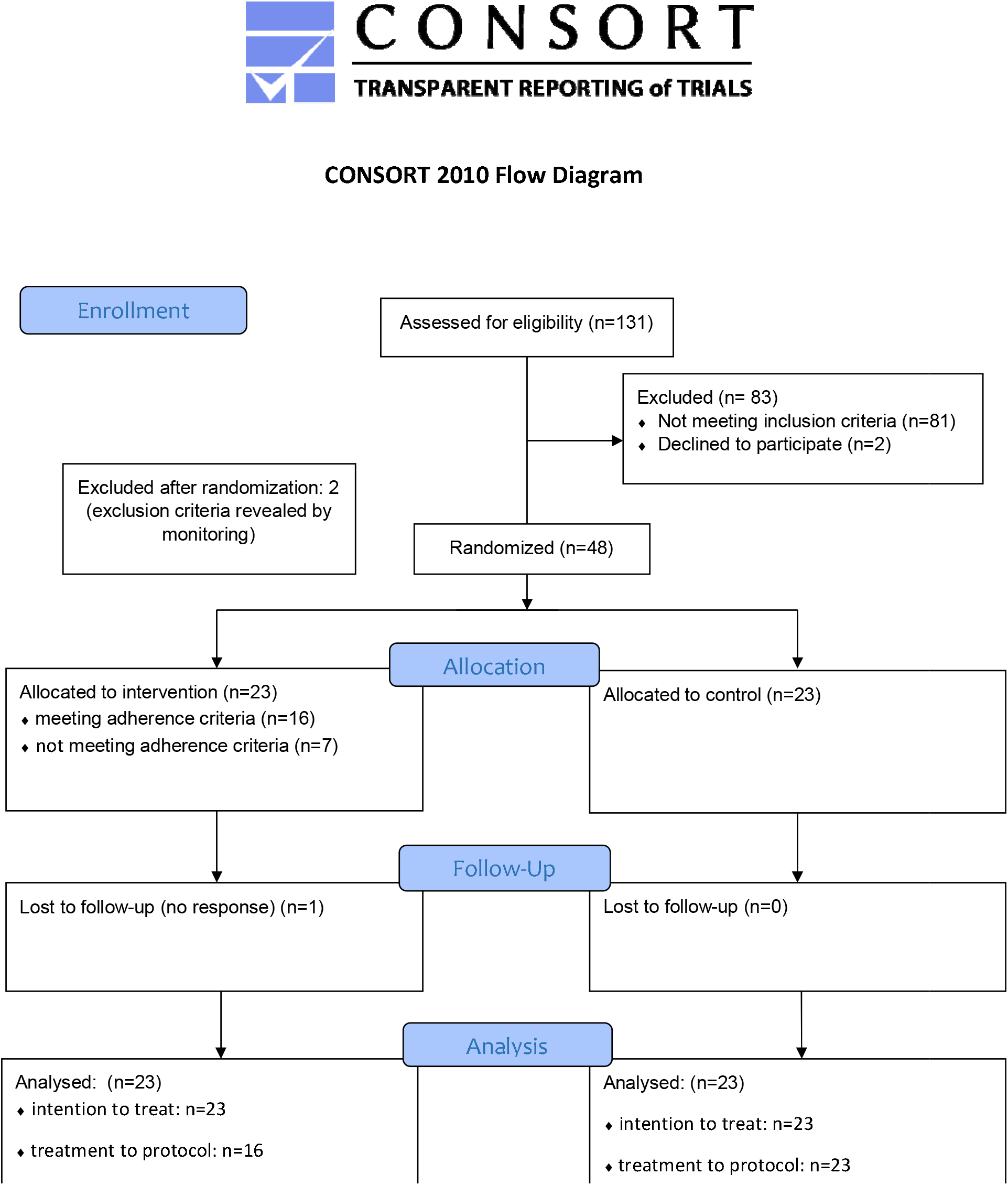
Flowchart Recruitment

Characteristics of the study samples are shown in Table 1. The study sample was between 23 and 83 years old (57 years±13 years), 60% were male. N=30 (65%) had relevant comorbidities. All patients showed pulmonary infiltrates in thoracic computer tomography. Mean hospitalization length was 9.8±3.1 days (range 5-19 days).

Although no statistically significant differences were found between intervention and control group for baseline variables, clinically relevant differences were apparent. Patients in the intervention group were older (M=58.8±13.2 vs. M=54.3±13.4), had more relevant comorbidities (N= 18, 78% vs. N=12, 52%) and a higher amount of COVID-19- and pneumonia-associated medication during hospital stay (M=2.8±1.0 vs. M=2.3±1.3) (see Table 1).

### Primary outcome

Estimated marginal mean course of log-transformed IL-6 for all patients of the ITT-sample (N=46) is shown in Figure 2A. The prediction models included the covariates set to mean (relevant comorbidities (no/yes), count of COVID-19 pneumonia medication, and age (years). Multilevel fixed-effect linear regression models were compared using likelihood-ratio tests (see Table 2). These LR tests identified a random slope model with a group by time interaction as the superior model for IL-6 (LR chi^2^ (13)=23.18; p=.040). The graphical results of the model (marginal means displayed in Figure 1) on average show lower values of IL-6 in the IG (effect size Cohens f^2^ =0.11, LR-test p=.040). Treatment-per-protocol analysis (N=39) confirmed these results (f^2^ =0.14, LR-test p=.022).

**Figure 2.**
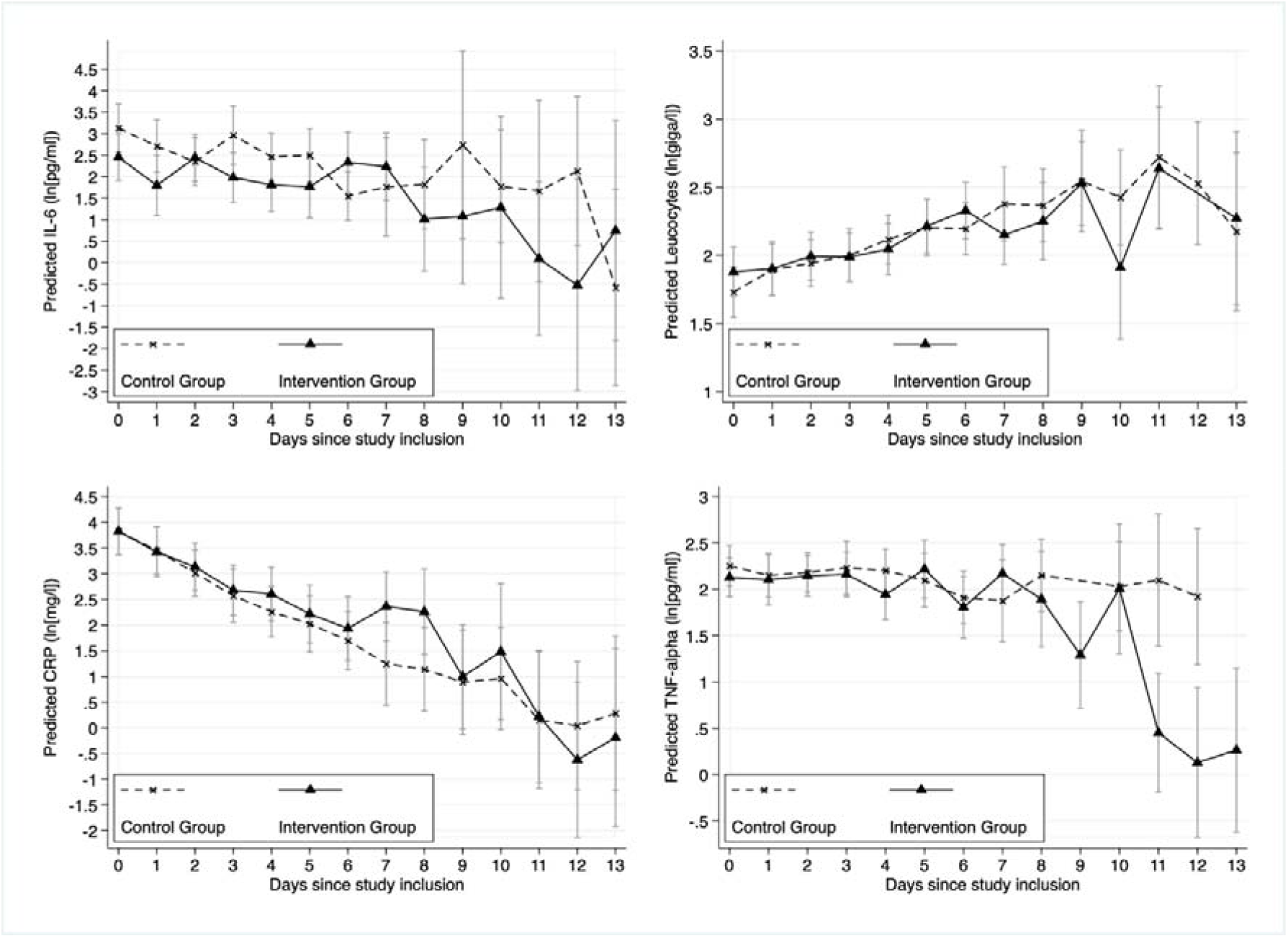
Trajectories of inflammatory outcomes: Marginal mean prediction of (A) ln(IL-6[pg/ml]), (B) ln(leucocytes[giga/l]), (C) ln(CRP[mg/l)] and (D) ln(TNF-α [pg/ml]) values for IG and CG from multilevel fixed-effect linear regression models with random slope (N=46 individuals with N=208 observations; average observations per individual=4.5). Note: Negative ln values translate to parameter values <1. Covariates: relevant comorbidities (no vs. yes), COVID-19 pneumonia medication (count), and age (years). Model predictions were calculated at covariate mean values.

### Secondary outcomes

Estimated marginal mean course of log-transformed leucocytes, CRP and TNF-α for all patients of the ITT sample (N=46) is shown in Figure 2B, C, D. The model comparison for the secondary outcomes leucocytes, CRP and TNF-α showed no relevant group by time interaction (see Table 2).

### Post hoc analyses

To further explore the relationship between slow-paced breathing and IL-6, we modeled a dose-response analysis using the daily minutes of slow-paced breathing to predict IL-6 of the next day. Marginal mean values from adjusted multilevel mixed-effect linear regression models are shown in Figure 3. The model indicates a dose-response relationship with beneficial effects of practice time above 45 minutes a day (b=-.82, 95%CI lower -1.55; upper -.01).

**Figure 3:**
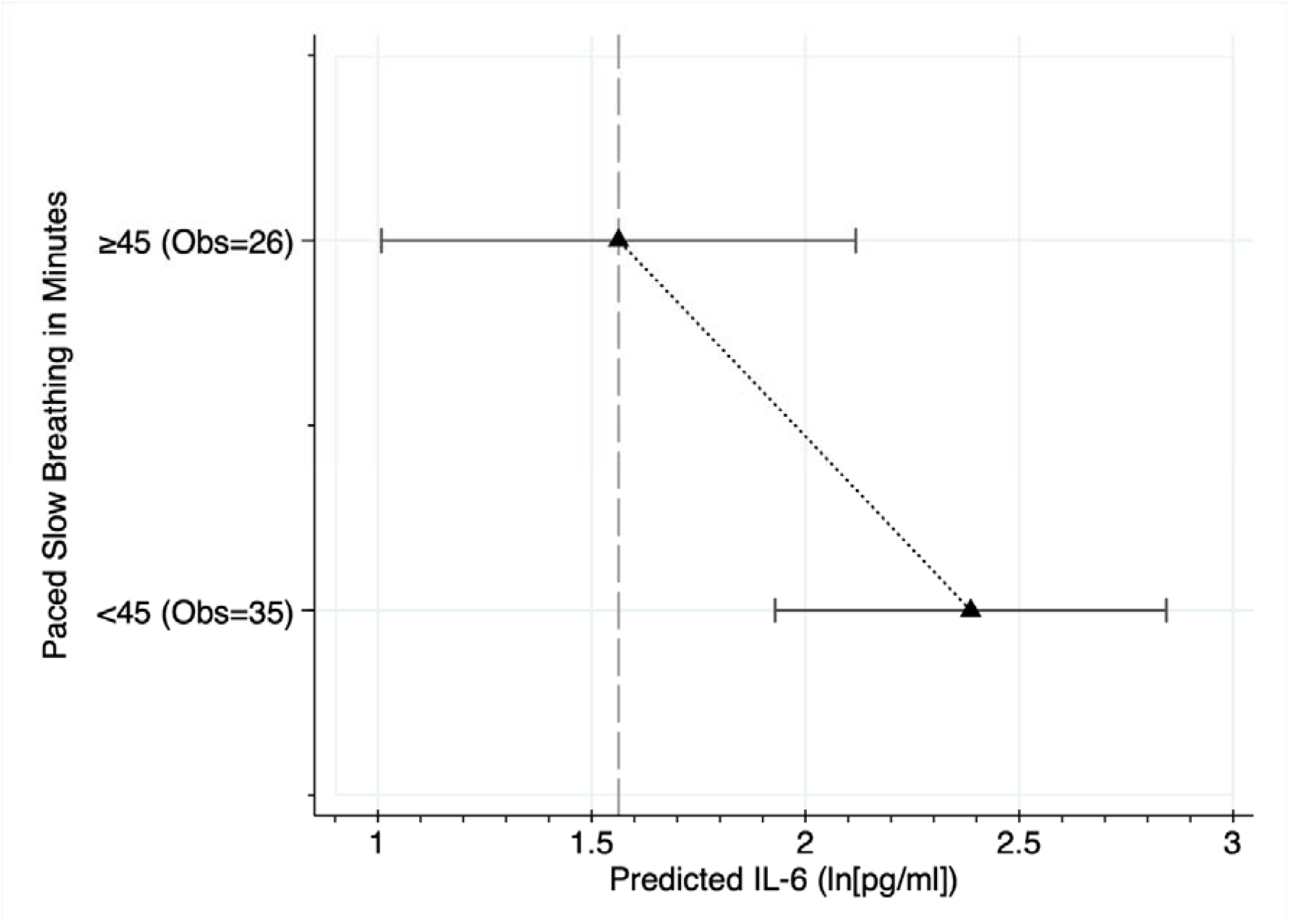
Dose response analysis using categorized breathing minutes from 22 patients and 61 days. Marginal mean values from multilevel mixed-effect linear regression models adjusted for relevant comorbidities (no vs. yes), COVID-19 pneumonia medication (count), and age (years). Prediction at covariate mean values. Obs. = Observations (patient days breathing)

### Adverse events

Six patients (N=3, 13 % in each study group) were admitted to the ICU, of whom one patient died (CG). All 23 patients practiced at least once. Most patients (N=18, 78%) managed the breathing exercise well. Nine patients (39%) reported the exercise at least from time to time as demanding. Two stopped due to difficulties with the implementation of the breathing exercise in terms of concentration and technique. There was one case of dizziness that resolved by reducing the depth of breathing slightly. Four out of the 23 patients (17%) who practiced at least once complained about coughing especially at the beginning of the exercise, and one had to stop the exercise once due to coughing. Weighted mean breathing frequency at rest was 18.5/min±4.5/min (range 10-30.5/min) for IG with no significant difference to the CG (M= 19.2/min±3.6/min, range 13.5-28.3/min; p=0.431. During the exercise, mean breathing frequency was 6.4/min±1.0/min (range 5.7-10/min) in IG. Six patients felt uncomfortable breathing at 6/min and had adjusted frequencies up to 10/min. 34 patients received nasal oxygen at least once during the hospital stay with a patient weighted mean flow of 1.9l ± 1.8l per min. Oxygen saturation was comparable between IG (95.1%±2.1%) and CG (94.7%±1.8%) at rest (p=0.444) but increased during slow-paced breathing (95.4%±1.6%, p=0.003).

## Discussion

This clinical trial of patients with moderate COVID-19 pneumonia showed that slow-paced breathing is effective to reduce IL-6 in COVID-19 pneumonia, though with uncertain clinical importance. Further, the data showed that reducing breathing frequency to 6/min is safe and feasible in moderate COVID-19 pneumonia and did not deteriorate oxygen saturation.

A non-invasive, non-pharmaceutical, not device dependent treatment option in COVID-19 disease has several advantages. It reduces the risk of toxicity of IL-6 receptor antagonists. Costs are low as no devices have to be bought or certified. The technique itself is easy to learn and the exercises can be supported via free apps on the patient’s own smartphone. Therefore, the intervention can be scaled easily by training medical assistance staff that instructs the breathing techniques and supports first practice sessions. This would offer an access to a therapy option not only for industrial, but also for low-income countries.

Our data adds to the knowledge about the effect of VNS on inflammation marker. To our knowledge, this is the first study showing a statistically significant direct effect of non-invasive VNS via paced-breathing on IL-6 of the following day. The exploratory dose-response analysis proposes a linear relationship with more minutes in slow-paced breathing reducing IL-6 values more the next day. The dose necessary for this effect was 45 minutes of paced breathing a day at a breathing frequency of 6/min with an inhalation to exhalation ratio of 4:6. Though data about the effect of slow breathing in acute inflammation is scarce, the data available for the effect of interventions including slow breathing on IL-6 seems to depend highly on the frequency and duration, with an effect only in studies with at least half an hour of practice daily (23,24). More detailed studies should further explore the necessary frequency, ratio and dose for a meaningful reduction in IL-6.

We cannot distinguish effects of VNS and placebo. Psychosocial interventions have been shown to affect the immune system (25). Though the amount of attention by study personnel was approximately the same in both groups, patients in the IG might have felt more self-efficient and this might have influenced their inflammatory marker. Though this effect would not be triggered by VNS, it would still originate in the central autonomic network and still, the anti-inflammatory pathway would be triggered drug- and device-free. From a patient’s point of view, this is very important. We had a very high rate of patients willing to participate (97.6%) because most patients were very interested in a study that investigates a therapeutic approach without drugs and devices. Furthermore, most patients were very happy to perform the intervention because it was their only possibility to manage their disease. This alleviated their feeling of being helpless and without control, introducing the feeling of self-efficacy.

### Limitations

The first limitation is the sample size that could not address clinical outcomes. Second, early discharge was not included in the model. Third, the intervention was not blinded. Another trial could include a sham intervention. Fourth, we did not control objectively the amount of time spent in slow-paced breathing but relied on self-report.

## Conclusion

This small, single-center randomized controlled clinical trial showed that reducing breathing frequency to 6/min is effective in reducing IL-6 levels in moderate COVID-19 pneumonia without relevant side effects. Larger RCTs need to confirm these results as well as evaluate clinical outcomes. This would offer access to a therapy option not only for industrial, but also for low-income countries.

## Supporting information

Consort Checklist

Study Personal Instruction Breathing intervention

## Data Availability

All data produced in the present study are available upon reasonable request to the corresponding author

## Author Contribution

Conceptualization: MNJ, EMB

Data curation: MNJ, EMB

Formal Analysis: MNJ, EMB

Investigation: EMB, BG, SH, MKG

Methodology: MNJ, EMB, JFT

Project administration: EMB, MNJ, BG

Resources: HG, BG

Software: EMB, MNJ, SH, MKG

Validation: EMB, MNJ, SH, MKG

Visualization: MNJ, EB

Writing – original draft: EMB, MNJ

Writing – review & editing: EMB, BG, SH, MKG, JFT, HG, MNJ

## Acknowledgements

We thank the directors of the Clinic for Internal Medicine III, Prof. Hartmut Döhner, and Clinic for Internal Medicine I, Prof. Thomas Seufferlein, for their support in conducting this study. Special thanks to Andreas Binzberger, Jinny Scheffler, the ward physicians in charge, who supported us during study conduction. Thanks to Michael Holl, developer of the BreathBall App, for the inspiring interaction. The authors declare no competing financial interests.

