## Supplementary material for "A randomized clinical trial to stimulate the cholinergic anti-inflammatory pathway in patients with moderate COVID-19-pneumonia using a slow-paced breathing technique": Study Personal Instruction Breathing intervention

### Instruction Slow-Paced Breathing Exercise

Our breathing and heartbeat are closely related to each other. When we inhale, our heart beats faster. When we exhale, it beats slower. This connection is based on your autonomic nervous system. This system controls a number of other important body functions, such as your blood pressure and immune system. Your autonomic nervous system is guided by your subconscious, but you can specifically influence the activity of your autonomic nervous system through your breathing - and by doing that also influence your immune system in addition to your heartbeat.

The effects on your heart can be determined by means of measurements, e.g., the fluctuation of your pulse, the so-called heart rate variability. This measurement is done using a simple chest strap that records the electrical signal from the heart muscle. By taking blood samples we measure the influence on the immune system. We want to investigate to what extent breathing exercises can improve heart rate variability and inflammatory markers.

In order to achieve an effect, it is important that you do the breathing exercise often enough and for a long time. **You should practice three times a day for 20 minutes.** However, it is also important that you feel comfortable doing the exercise. No effect is achieved under excitement or tension. If you do the exercise correctly, you will always feel more relaxed. You may become tired or even fall asleep. This is proof that you have performed the exercise correctly.

If you feel uncomfortable during the exercise or your fingers start tingling, you get dizzy or show signs of headache and nausea then something is wrong and you should stop the exercise and contact the study staff. They will check with you whether you need to improve your technique or rather stop doing the exercise.

The BreathBall app will help you doing the exercise on your own (Setting 4:6 Inhale:Exhale ratio)

**Brief instruction (by study personnel):**

- When the breath ball gets bigger, you should inhale.
- When it gets smaller, you should exhale.
- Here you can start the exercise, here you can stop it, here you can adjust the speed.
- Once again, it is very important that you breathe calmly and in a relaxed manner. Do not overexert yourself. If you feel dizzy, try to breathe more shallowly.
- Now go ahead and try Breath Ball.
- Be sure to breathe into your belly. You can put one hand on your belly and one on your chest in order to get a better feeling for the exercise. The hand on your belly should move more. Like this: (show).
- While doing this, you can also imagine breathing in all the way down to your feet.
- Slightly pressing your lips together while exhaling (lip brake) can also help.
